# Integration of metabolomic and genetic data reveals novel variants underpinning the human metabolome: the Coronary Artery Risk Development in Young Adults (CARDIA) study

**DOI:** 10.64898/2026.01.28.26344965

**Authors:** Mohanraj Krishnan, Blake R. Rushing, Annie Green Howard, Heather M. Highland, Victoria L. Buchanan, Samson Okello, Yao Tu, Shuwei Liu, Xiuxia Du, Susan L. McRitchie, Myriam Fornage, Cora E Lewis, Jared P Reis, Susan J. Sumner, Christy L. Avery, Penny Gordon-Larsen, Mariaelisa Graff, Kari E. North

## Abstract

Metabolite genome-wide association studies have identified hundreds of variants, many of which play intermediate roles linking genotype and phenotype with downstream diseases. However, the majority of metabolite GWAS have been published in self-identified Non-Hispanic White (NHW) populations, greatly limiting inference to other populations. Here we report the results of a GWAS of 7,522 untargeted metabolite peaks in 2,183 participants of the Coronary Artery Risk Development in Young Adults (CARDIA) study (714 Black and 1,469 White individuals, mean age = 43.56, women = 56.1%). We performed untargeted metabolomic profiling on plasma samples from these individuals using ultra-high performance liquid chromatography-high resolution mass spectrometry. Using race/ethnicity-stratified and combined GWAS of 8,534,915 (Black stratum) and 5,886,255 (White stratum) common variants (minor allele frequency ≥ 0.05) from TOPMed Imputation Reference Panel r^2^, we examined association with the CARDIA 7,522 metabolite peaks. We used MetaboAnalyst 5.0 and publicly available phenome-wide association studies to predict functional pathway activity and characterize unknown peaks. We identified 171 significant (*P*-value < 6.6×10^-13^) sentinel variants across 536 metabolite peaks, of which 39 were annotated with our in-house library in the race-combined CARDIA analysis. PheWAS and pathway enrichment models provided information on unannotated metabolite peaks including the novel *KHNYN/SDR39U1* locus which supports vitamin B5’s involvement in the citric acid cycle. We also identified a race-specific variant (rs79530723) near *SLC28A1*, which plays an important role in cytidine metabolism and was specific to the CARDIA Black participants. Pathway analyses prioritized a role of pyrimidine metabolism for this peak, a finding supported by studies in the Hispanic/Latino populations. Overall, our study results support a strong shared genetic architecture of the human metabolome across diverse populations and reveal new metabolite GWAS signals, underscoring the value of integrating ‘omics’ techniques to enhance comprehension of its genetic architecture.

## BACKGROUND

Metabolites are small molecules (<1.5kDa) (1) that are the precursors, intermediates, co-factors, and end-products of cellular metabolism (2, 3). Variation across the human metabolome reflects homeostatic dysregulation and provides a window into biological mechanisms of health and disease (4). The coupling of genome-wide association analysis (GWAS) with metabolomic profiling has successfully identified hundreds of variants associated with a broad range of metabolites, many of which also associate with complex diseases (1, 5–18). These metabolomic GWASs provide novel insights into the potential molecular mechanisms associated with metabolic diseases (6, 19, 20). Despite these advances, the majority of mGWAS have been conducted in Non-Hispanic White populations (NHW) (15, 18), limiting inference to only a fraction of the world’s population (21). Only a handful of mGWAS have been performed in non-White populations (1, 15, 18), all of which have identified population-specific genetic determinants of circulating metabolites. Integrative analyses of genomics and metabolomics are increasingly being used to characterize unannotated metabolites and their relationship to downstream disease, and it is critical that all populations benefit from these studies to ensure equitable precision medicine advances.

Here, we performed a GWAS of circulating metabolites from an untargeted metabolomic panel of 7,522 metabolomic peaks in 714 participants of Black race and 1,469 participants of White race from the Coronary Artery Risk Development in Young Adults (CARDIA) study. We implemented pathway enrichment analysis and interrogated publicly available phenome-wide association studies (PheWAS) to reveal relationships between genetics, metabolites, and underlying biological processes. These findings complement the catalog of genetic loci associated with metabolites and may provide potential targets for intervention especially among populations that are most burdened with disease and underrepresented in scientific research.

## METHODS

### Study design

The Coronary Artery Risk Development in Young Adults (CARDIA) cohort is a multi-center, prospective, community-based study conducted in four US metropolitan areas (Birmingham, AL; Chicago, IL; Minneapolis, MN; and Oakland, CA) (22) investigating the origins of cardiovascular disease. The study began in 1985 (exam year 0) with 5,115 men and women of self-identified Black or White race aged 18 to 30 years. At each site, the CARDIA sample was designed to comprise nearly equal numbers of participants by sex, self-identified race, age (18–24 years or 25–30 years), and education (less than high school, or high school or greater). Of 3,549 participants at the year 20 CARDIA exam, we generated untargeted metabolomics in 3,391 participants. We excluded participants if they did not meet quality control and pre-processing measures (N=31), were missing (>50%) metabolite (N=3), were pregnant (N=6), had mismatched gender (N=1) or did not consent to genetic testing (N=1,167) (**Figure 1**). This resulted in a final analytic dataset of 2,183 participants of whom 714 were Black (female = 63%) and 1,469 were White (female = 53%) (23). The protocol was approved by the institutional review boards of each participating institution; each study participant provided written informed consent.

**Figure 1:**
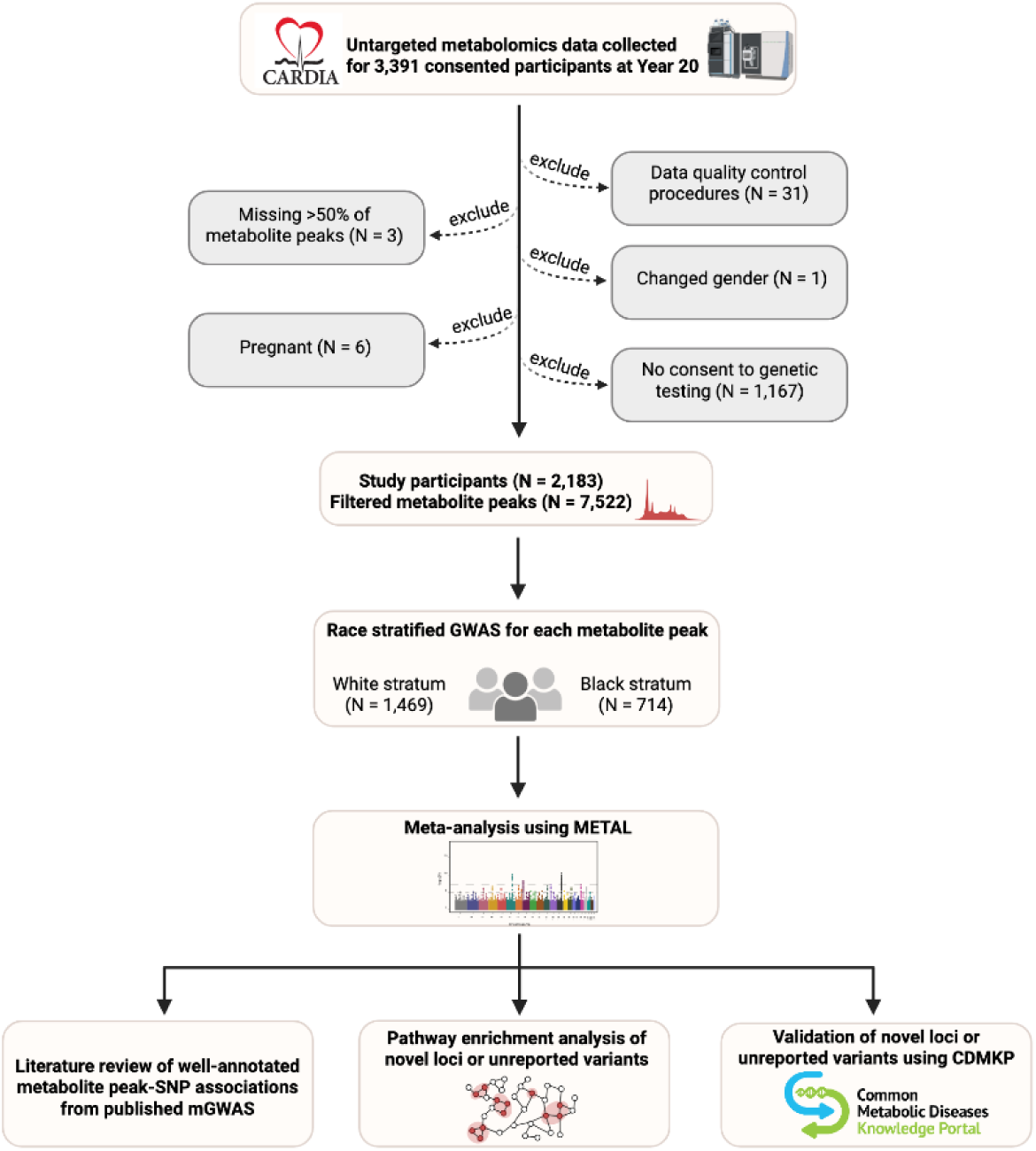
Schematic of the study design for genomic analysis and downstream systematic analysis of metabolite peaks in CARDIA.

### Metabolomic profiling

Metabolomic profiling was conducted using stored frozen fasting plasma samples (50µL) from the Year 20 CARDIA exam using a Vanquish ultra-high performance liquid chromatography (UHPLC) Q Exactive^TM^ HF-X Hybrid Quadruple-Orbitrap^TM^ Mass Spectrometer (Thermo Fisher Scientific, San Jose, CA, USA). Samples were randomized across 42 batches by sex, race, and 10-year atherosclerotic cardiovascular disease (ASCVD) risk. Quality control study pools (QCSPs) were created for each sample batch by combining an additional 10µL of each study sample into a singular mixture and aliquoting into tubes at 50µL each. Method blanks were created by aliquoting Liquid Chromatography-Mass Spectrometry (LC-MS) grade water into tubes at 50µL each. External reference material was prepared by aliquoting National Institute of Standards and Technology (NIST) reference plasma (SRM 1950) into tubes at 50µL each. All study samples, QCSPs, blanks, and NIST samples were processed identically. QCSPs, blanks, and NIST samples were interspersed at a rate of 10% throughout the study samples for each batch during data acquisition. Raw metabolite data was preprocessed using Automated Data Analysis Pipeline (ADAP) (24). Data were normalized to total intensity and batch effects were corrected by filtering LC-MS peaks in NIST samples that had an Analysis of Variance (ANOVA) *P*<0.05 across the 42 batches, leaving a total of 7,522 metabolite peaks for analysis (**Supplementary Table 1**). The resulting metabolite peaks were median scaled and missing data for metabolite peaks were imputed using random forest (which has shown to perform well using untargeted metabolomics data) (23, 25). Median-scaled and imputed data were then log_2_ transformed to ensure normal distribution of each metabolite across samples. All peaks were input into the Automated Data Analysis Pipeline Spectral Knowledgebase (ADAP-KDB) (26) to match peaks to metabolites resulting in 689 peaks matching to 354 unique metabolites in the in-house physical standards library of retention time (RT), mass spectrometry (MS), and tandem mass spectrometry (MS/MS) fragmentation.

Additional matching was performed using public databases (NIST, Human Metabolome Database (HMDB)) via MS and or experimental MS/MS. An ontology system was used to denote the evidence-basis supporting each match (27, 28) In addition, mass, and retention time values for all 7,522 peaks were input into the “Functional Analysis” module of MetaboAnalyst 5.0 and matched to metabolites in the Kyoto Encyclopedia of Genes and Genomes (KEGG) database using the *mummichog* algorithm for pathway analysis and additional metabolite annotation. The *mummichog* algorithm is an algorithm designed to identify significant pathways using unannotated mass spectral peaks using mass and retention time data and matching metabolite databases. A mass tolerance of 5ppm was used, and primary ions were not enforced. All candidate matches derived from mummichog results were recorded for each peak.

### Genotyping and Imputation

Genotyping in CARDIA was performed through the National Heart, Lung, and Blood Institute (NHLBI) Candidate Gene Association Resource (CARe) consortium using the Affymetrix Genome-Wide Human 6.0 array (29, 30). Haplotype-based imputation was performed using phased haplotypes from TOPMed r^2^ as reference using the TOPMed imputation server (31). Post-imputation quality filtering was performed using a R^2^ threshold specific to each minor allele frequency (MAF) category to ensure average R^2^ for variants passing threshold was at least 0.8 (32).

### Metabolite Associations with GWAS imputed variants

For each metabolite peak, we regressed out the covariate effects of age at year 20, sex, and metabolomic batch, and inverse rank normalized the residuals. These residuals were used as the dependent variable in the GWAS analysis. To allow for differences in LD across populations we first performed race stratified GWAS of 8,534,915 (Black stratum) and 5,886,255 (White stratum) imputed genetic variants MAF >0.05, imputation r^2^ >0.3) across normalized residual metabolite values adjusting for the first ten ancestral principal components (PC) using Plink 2.0 (33, 34), followed by a joint meta-analysis (via inverse-variance method) using METAL (35).

Given the continuous nature of ancestral variation, our primary analysis was the meta-analysis. However, we also considered race-stratified analysis to query unique race-specific variants. Significant single-nucleotide polymorphism (SNP)-metabolite peak associations were identified according to a Bonferroni adjusted *P* value of 6.6×10^-13^ (5×10^-9^/7,522 peaks) within 1MB windows using the *topr* package (36). Stepwise conditional analysis was performed for metabolite peaks with a genome-wide significant SNP in each race group using the conditional and joint (COJO) Genome-wide Complex Trait Analysis (GCTA) software package to identify near independent association signals (37). For metabolites with statistically significant estimated SNP effects, we first conditioned on the associated variant of the highest statistical significance and continued conditioning on the most significant remaining variant until no variant attained a significance of 6.6×10^-13^. Annotation of SNP-metabolite associations were conducted using the Ensembl Variant Effect Predictor (VEP) (38). To annotate the identified locus-metabolite associations, we queried the GWAS Catalog, PhenoScanner v2.0 and performed manual searches of the literature (1, 15, 17) to contextualize our findings. If a variant belonging to a region-metabolite pair was reported to be associated with any of the metabolites, the region-metabolite pair was considered known, otherwise it was considered unreported in the literature.

### Pathway enrichment analysis

Pathway enrichment analysis was performed using the *mummichog* algorithm (version: 2.3.3-20200213, default metabolic human model (KEGG) implemented in MetaboAnalyst 5.0) (39). Mummichog is a bioinformatics Python-based platform that infers and categorizes functional biological activity using direct peak outputs from liquid chromatography-mass spectrometry instruments, including mass-to-charge (m/z) and retention time (r/t). The algorithm searches tentative compound lists from metabolite reference databases against an integrated model of human metabolism to identify functional activity. Fisher’s exact tests (FET), adjusted for type I error through a pathway permutation procedure, were used to calculate *P* values for significance at the pathway level. Specifically, the likelihood of pathway enrichment across significant features is compared to pathways identified across the entire KEGG compound list, considering the probability of mapping the significant metabolic features to pathways. *Mummichog* parameters (m/z ratios and *P* values) from both our joint and stratified analyses were set to match against ions included in the ‘positive mode’ setting at ± 5ppm without enforcing primary ions. Using MS peaks functional analysis the *P* value cutoff value of the metabolic pathway influence was set to 0.05 and pathways with a FET value less than 0.05 were selected as potential key metabolic pathways.

### Human genetic association validation using the Common Metabolic Disease Knowledge Portal (CMDKP)

Statistically significant (at Bonferroni corrected *P* value of 6.6×10^-13^) locus-metabolite peak association results were input into the Accelerating Medicines Partnership-Common Metabolic Diseases Knowledge Portal (AMP-CMDKP; cmdkp.org), a public resource that aggregates genetic association results (Human Genetics Knowledge Portal - Home. https://hugeamp.org/) from 461 datasets consisting of 488 phenotypes representing cohorts from 66 countries (40). We tested the identified variant around the genetic region of interest (region defined as the coding sequence of the variant plus 50kb of upstream and downstream flanking sequences) and variant level associations, which produce PheWAS plots for 20 different trait groups. We also checked effector gene predictions which integrated disease relevant data to predict likely causal genes at associated loci.

## RESULTS

Following preprocessing raw metabolomics data, including background filtering and batch effect corrections, 7,522 metabolite peaks remained in the CARDIA dataset for downstream GWAS. Principal component analysis showed tight clustering of QCSPs, blanks, and external reference materials, as well as acceptable centering of QCSPs within the study samples, indicating high data quality according to metabolomics standards (**Figure S1**) (41). The 7,522 metabolite peaks were then matched to compounds in either the in-house chemical standard library or to compounds in public databases (**Table S1**). While all association analyses used all 7,522 peaks, our interpretation of gene-metabolite linkages focused primarily on the in-house library matches due to the higher evidence basis supporting these matches.

### GWAS summary of the 7,522 metabolomic peaks in unstratified and race-stratified CARDIA participants

In the meta-analyzed CARDIA sample, we identified 171 sentinel variants across 536 metabolite peaks of which 39 metabolites were annotated with our in-house library standard (**Figure 2A**, **Figure 2B, Table S2**). In the CARDIA Black stratum, we mapped 31 sentinel variants across 100 metabolite peaks of which 12 metabolite peaks were matched to the in-house library (**Figure 2B, Table S3**). In the CARDIA White stratum, we mapped 148 sentinel variants across 448 metabolite peaks of which 32 peaks were matched to in-house library standards (**Figure 2B, Table S4)**.

**Figure 2.**
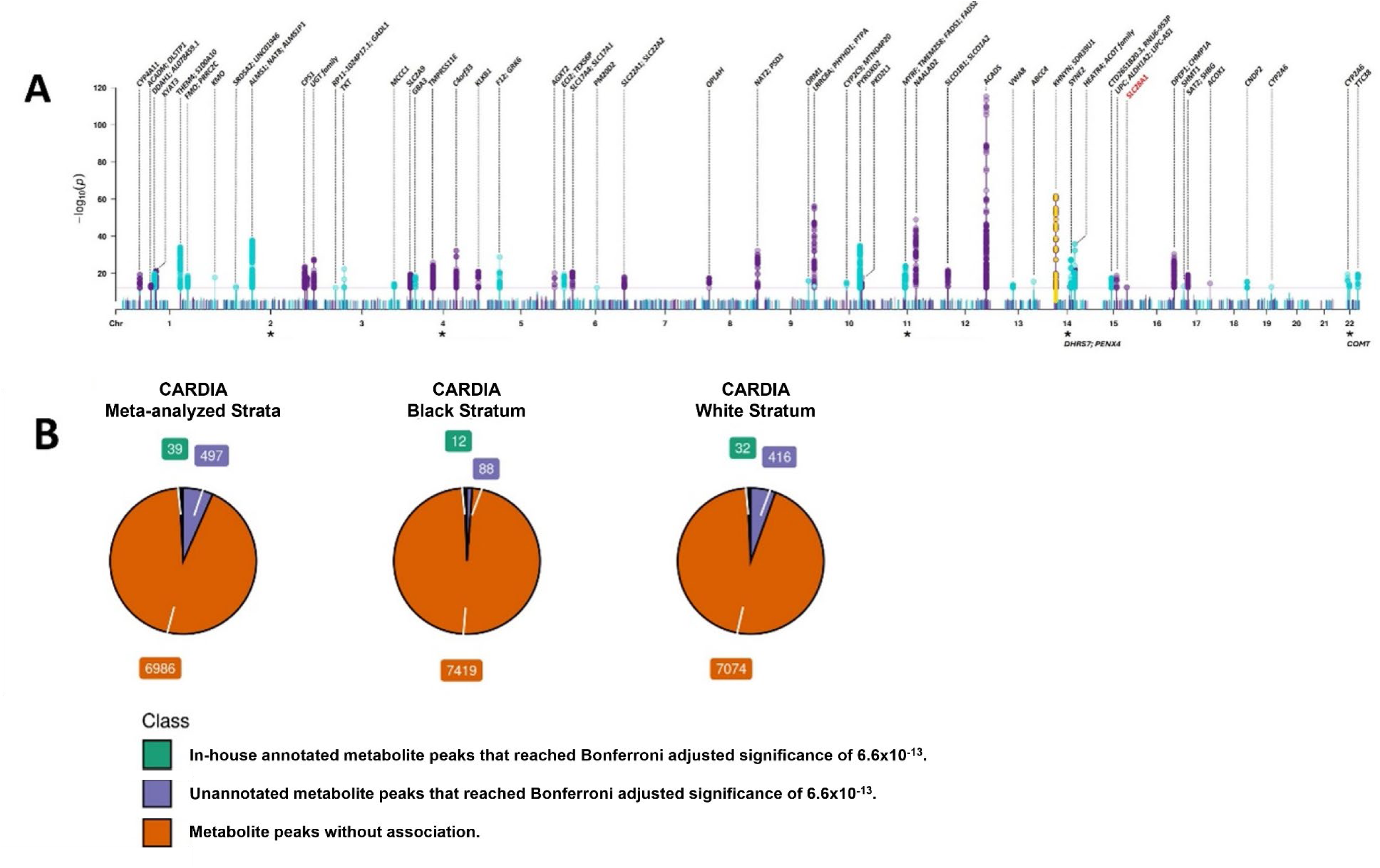
**A) Manhattan plot summarizing significant locus-metabolite peak associations in our CARDIA meta-analyzed and race-stratified strata.** Novel locus-metabolite peak association that do not overlap with those identified in previous large-scale metabolomic GWAS is shown in yellow. Known locus-metabolite peak associations are shown in blue and purple. Population-specific associations in the CARDIA Black stratum is labelled in red. **Figure 2B) Classification of tested metabolite peaks with or without genetic associations in the CARDIA meta-analyzed and race stratified strata.** Annotated metabolites are peaks that match to a standard in the in-house library of retention time (RT), mass spectrometry (MS), and tandem mass spectrometry (MS/MS) fragmentation. Unannotated metabolites are peaks that have an association but has no match with the standard in-house library.

#### Detecting independent association signals in our CARDIA race-stratified stratum

Conditional analysis using COJO (37) was only conducted in our race-stratified (Black or White) CARDIA stratum due to differences in LD patterns across populations. We found no secondary association signals across the Black stratum; however, seven metabolite peaks showed significant independent secondary associations in the White stratum with many of them occurring in the *DDAH1* locus. **Table S5** and **Table S6** display the variants for each metabolite peak that met our Bonferroni adjusted *P*-value (6.6×10^-13^) after conditional analysis.

#### Novel association between *KHNYN/SDR39U1* locus with an unannotated metabolite peak

New locus-metabolite peak associations in our study were vetted using PhenoScanner V_2_ (42), the GWAS Catalog (43), the “Every Gene Ever Annotated” public repository collected by Eric Fauman (17, 44, 45), as well as manual review of GWAS in diverse study populations, (Feofanova *et al* 2020 (1), Tahir *et al* 2022 (15), Yousri *et al* 2018 (18), Schlosser *et al* 2023 (46). We found a novel locus at the KH and NYN containing /Short Chain Dehydrogenase Reductase Family 39U Member 1 (KHNYN/*SDR39U1*) region to be associated with an unknown metabolite peak (m/z=176.1281, r/t=2.38 min). The sentinel variant rs11158703 in the meta-analyzed CARDIA analysis (splice region: c.328+4C>T, [MAF=0.213], β=-0.658, SE=0.04, *P*=1.61 ×10^-65^) (**Figure 3A**), along with two other markers in high LD; rs4981516 (intergenic, [MAF=0.462]) in the CARDIA Black stratum and rs7151530 (intronic: n.280+7910G>T, [MAF=0.091]) in the CARDIA White stratum, were associated with the same unknown metabolite peak [m/z = 176.1281, r/t = 2.38 min] (**Figure S2**). Functional analysis matching metabolites in the KEGG database, pathway enrichment analyses and curated PheWAS were conducted to predict pathway level effects and to functionally characterize the association. Matching based on m/z ratio and retention time by the *mummichog* algorithm in MetaboAnalyst 5.0 characterized this metabolite peak as pantothenic acid (Vitamin B5). Pathway enrichment analysis using all 7,522 metabolite peaks and their respective association *P* values identified a total of five, seven, and five statistically significant pathways (FET <0.05), across the meta-analyzed (**Figure 3B**), Black and White strata (**Figure S2**) respectively, with the citric acid cycle (TCA) being common across all three analyses. PheWAS, using the AMP-CMDPK public-based forum, also showed some enrichment of rs11158703 with citrate (β=-0065, *P=*0.015) (**Figure 3C**). Leveraging raw spectral metabolomic profiles, pathway enrichment and publicly available PheWAS, we hypothesized that the metabolite peak, which did not have any matches to the in-house library (pk11747), was a compound that was important within the citric acid cycle, most likely a derivative of pantothenic acid. This potentially adds more clarity to the identity of this peak which had 90 potential matches at a pd_d level (mass match only) to public databases (HMDB and NIST).

**Figure 3:**
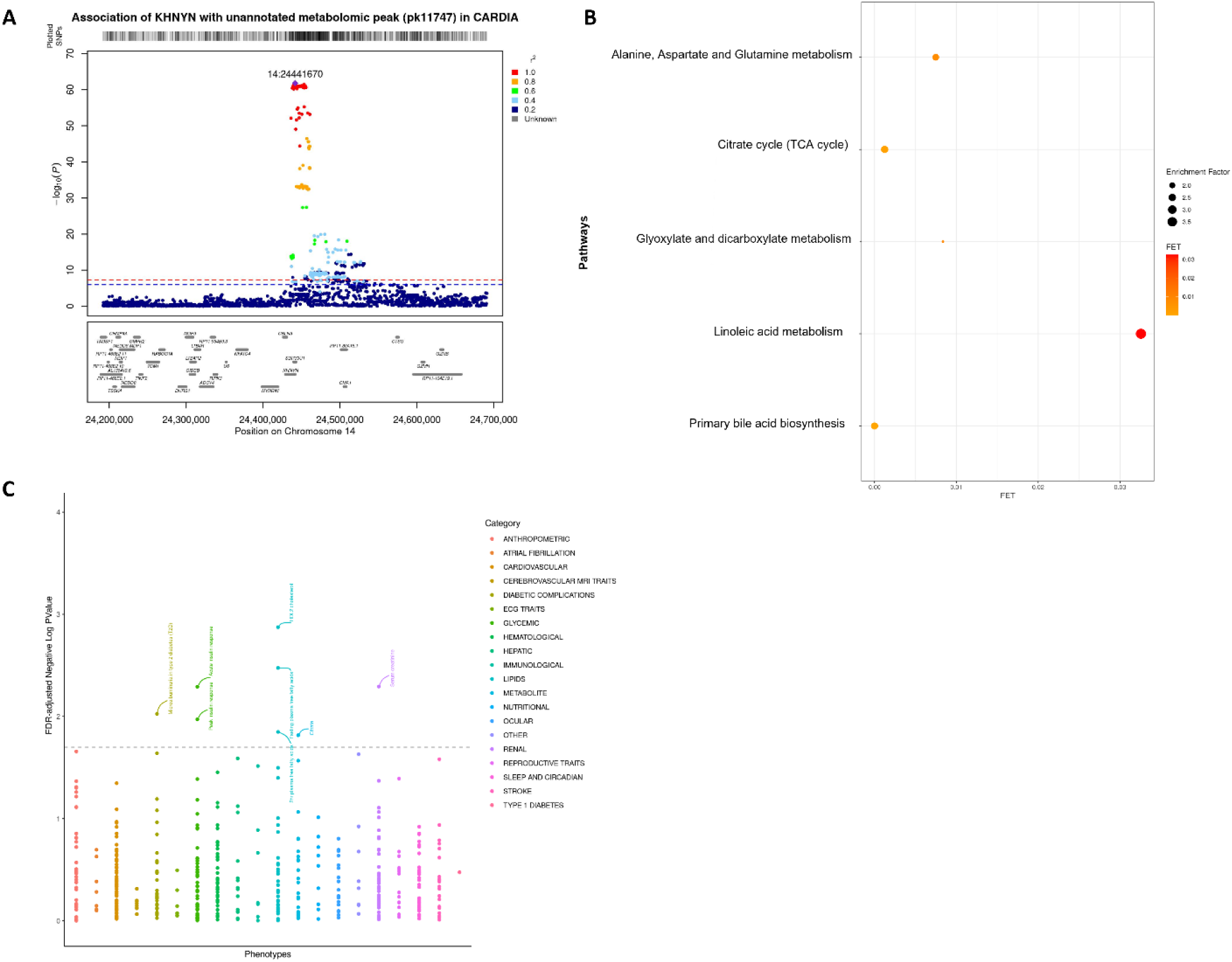
**A: Regional association (LocusZoom) plot from the region encompassing *KHNYN/SDR39U1* in the meta-analyzed CARDIA sample.** The top variant (rs11158703) is indicated with a purple diamond. The red line corresponds to a *P* of 6.6 ×10^-13^. The blue line corresponds to a *P* of 5 ×10^-8^. **B: Dot plot of statistically significant pathways (<FET 0.05) from functional enrichment analysis of the CARDIA 7,522-metabolomic peaks using *P* values corresponding to rs11158703 and m/z ratios for each peak.** Each dot represents a metabolic pathway and is plotted according to its Fisher’s exact test (FET) *p*-value on the x-axis. The dot size indicates the size of the enrichment factor, which represents the ratio of significant to total expected hits for each pathway. **C: PheWAS results for rs11158703** from the Common Metabolic Disease Knowledge Portal from https://hugeamp.org.

#### Directional consistency in metabolite-SNP associations in CARDIA Black and White stratum

Of the 100 significantly mapped SNP-metabolite peak associations across CARDIA Black stratum, 75% of the associations showed consistent direction of among CARDIA White stratum (*P=*1.81×10^-46^, *R_correlation_*=0.97[0.95∼0.98]) (**Figure S3A Table S7**). Similarly, 91.1% of the SNP-metabolite peak associations identified in the CARDIA White stratum showed a consistent direction of effect in the CARDIA Black stratum (*P=*21.52×10^-156^, *R_correlation_*=0.91[0.89∼0.92]), (**Figure S3B, Table S8**). The remaining variants that were missing were either removed prior to GWAS analysis due to poor imputation quality, were population-specific variants, and/or the MAF was <5% in that respective stratum.

#### Evidence for known and novel variants in established loci with annotated metabolomic peaks

Overall, most of our annotated single-metabolite GWAS associations were consistent at the metabolic pathway level, supporting a shared genetic architecture of the human metabolome. Of the 33 in-house library metabolite-SNP associations in the meta-analyzed samples (excluding duplicate SNP-metabolite associations from different metabolite peaks), 11 variants (33%) were mapped to previous SNP-metabolite associations from published metabolomic GWAS (**Table S9**) (1, 15, 17, 18, 44–46). Sixteen (48%) reported variants (1, 15, 17, 18, 44–46) were associated with new metabolites and two unreported variants [rs72552763 and rs4924750] in *SLC22A1* and *SHMT1* were associated with Butanoylcarnitine and Cotinine, respectively. Of the nine metabolite peak-SNP associations in the CARDIA Black stratum, five variants (56%) were mapped to known metabolites, three known variants (33%) were associated with new metabolites (1, 15, 17, 18, 44–46), and one novel variant near *PYROXD2* (rs59318566) was associated with L-Carnitine. In the CARDIA White stratum, 12 SNP-metabolite peak associations were consistent with known metabolites, 18 known variants (56%) were associated with new metabolites (1, 15, 17, 18, 44–46), and two novel variants [rs34142813 and rs4924750] located in *ALMS1* and *SHMT1* were associated with N-Octanoylglycine and Cotinine respectively.

#### Variants unique in race-stratified analyses

We found three sentinel variants (rs466067, rs558946049 and rs79530723) in the CARDIA Black stratum that were nearly monoallelic in individuals of non-Finnish European (NFE) ancestry (MAF<0.02% in gnomAD v4.0.0) and were not typed in our CARDIA White stratum. The African-specific variants rs466067 in *AGXT2* (synonymous: p.Gly435Gly, [MAF=0.123], β=-0.615, SE=0.08, *P*=7.02 ×10^-15^), and rs558946049 in *SLC22A24* (intronic: c.831-5865A>G, [MAF=0.065], β=0.834, SE=0.107, *P*=7.98 ×10^-15^), were associated with two unannotated metabolite peaks in the CARDIA Black stratum. However, the variants rs40199 in *AGXT2* (intronic: c.363-511C>T, [MAF=0.102], β=0.520, SE=0.06, *P*=9.81 ×10^-17^) and rs1939769 in *SLC22A24* (intronic: n.13-7777A>G, [MAF=0.055], β=1.28, SE=0.08, *P*=1.49 ×10^-56^) were also mapped to the same unannotated metabolite peaks in the CARDIA White stratum. The LD R^2^ between rs466067 and rs40199 and rs558946049 and rs1939769 using LDlink (including only the African ancestry) (47) was 0.0552 and 0.0301 demonstrating independent SNP effects in the same locus across strata. Only, the rs79530723 variant near *SLC28A1* ([MAF=0.088], β=-0.673, SE=0.09, *P*=4.57 ×10^-13^) was exclusively mapped to an unannotated metabolite peak (m/z=2.69024, r/t=1.01 min) in our CARDIA Black stratum (**Figure 4A**). Pathway analysis identified enrichment of the rs79530723-*SLC28A1* metabolite-peak with nicotinate and pyrimidine metabolism (**Figure 4B**).

**Figure 4:**
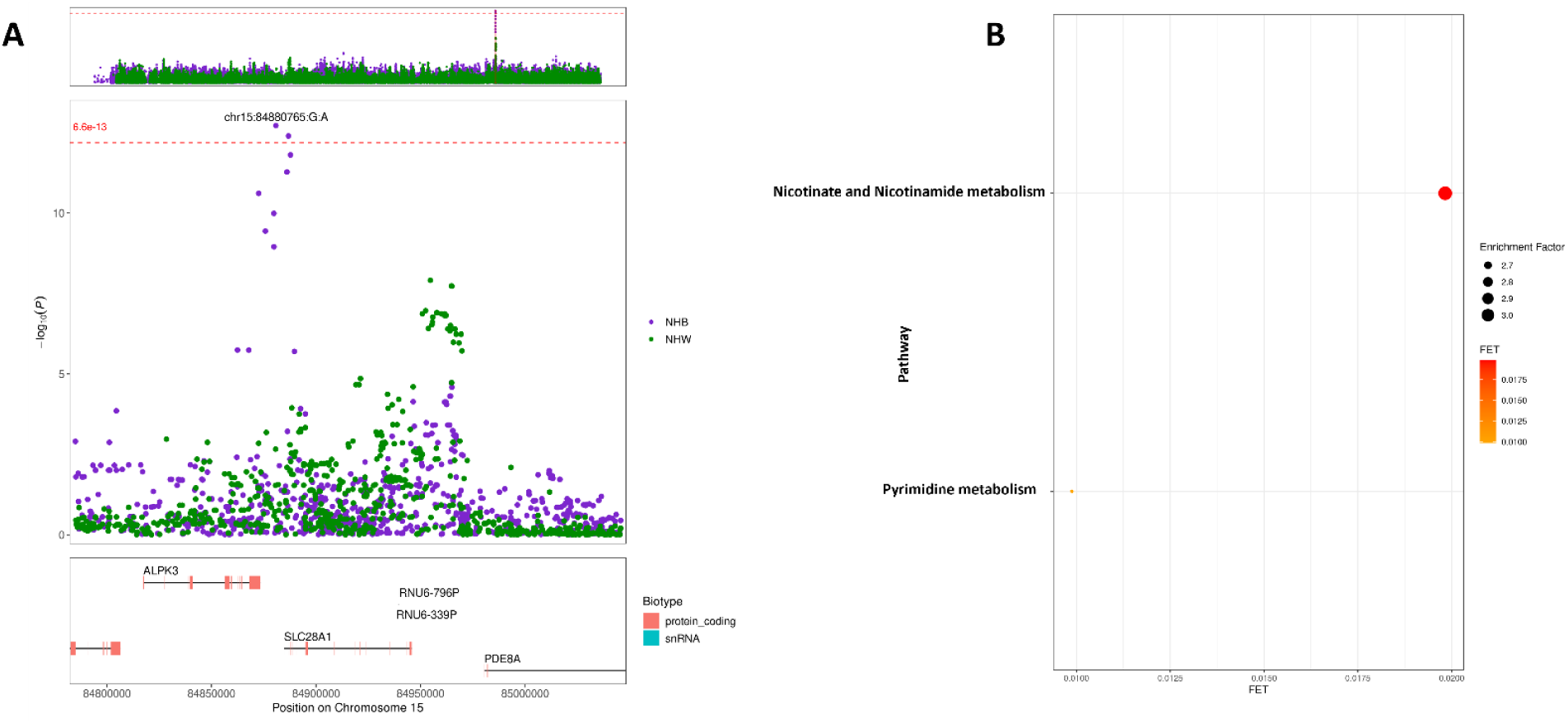
**A: Regional association plot from the joint analysis for the region encompassing *SLC28A1* in the race-stratified CARDIA Black and White stratum.** The top variant (chr15:84880765: G:A [rs79530723]) exclusively maps to the CARDIA Black stratum. The red line corresponds to a *P* value of 6.6 ×10^-13^. **B: Dot plot of statistically significant pathways (<FET 0.05) from functional enrichment analysis of the CARDIA 7,522-metabolomic peaks using *P* values corresponding to rs79530723 and *m/z* ratios for each peak from the CARDIA Black stratum.** Each dot represents a metabolic pathway and is plotted according to its Fisher’s exact test (FET) p-value on the x-axis. The dot size indicates the size of the enrichment factor, which represents the ratio of significant to total expected hits for each pathway.

## DISCUSSION

We performed GWAS to identify genetic signals for 7,522 metabolomic peaks among 2,183 (714 self-reported Black and 1,469 self-reported White) participants from the CARDIA study, allowing for differences in LD using race stratified GWAS of 8,534,915 (Black stratum) and 5,886,255 (White stratum) imputed genetic variants. We identified 536 metabolite peak-SNP associations in meta-analysis and 100 and 448 SNP-metabolite peak associations in CARDIA self-reported Black and White strata, respectively; many of which have been reported in the literature (1, 15, 17, 18, 44–46). We mapped one unreported locus-metabolite peak [*KHNYN*/*SDR39U1*] association in the meta-analyzed and stratified analyses and one SNP- (unannotated) metabolite association [near *SLC28A1*] in the CARDIA self-reported Black stratum. These findings add to the growing literature on the importance of genomic discovery across diverse study populations.

Early metabolomic GWAS (8, 48) have used older techniques to profile the human metabolome resulting in associations with large families of metabolites (49–51). Over time, optimized sample preparation procedures (52) along with enhanced profiling technologies (53) and automated data processing (54, 55) has allowed for wider metabolite coverage, better distinction of closely related compounds, and improved identification/annotation of individual peaks. Thus, newer genetic studies of the human metabolome will not only validate prior associations but reveal new associations that may have been previously missed. Here we found one novel association in the *KHNYN/SDR39U1* locus (rs11158703 in meta-analyzed, rs4981516 in the Black stratum and rs7151530 in the White stratum) with an unannotated metabolite peak; a likely derivative of pantothenic acid (Vitamin B5) based on raw mass spectral data. However, very little is known about the *SDR39U1* and the *KHNYN* genes, especially as it pertains to pantothenic acid. Previous studies reported SNPs in *KHNYN that* were associated with known metabolic traits, including height (56), blood (57), and brain phenotypes (58, 59).

In addition, *KHNYN* is an essential zinc finger antiviral protein (*ZAP*) cofactor that inhibits HIV-1 gene expression (60) and the citric acid cycle pathway has been shown to be important in responding to viral infections (61). Pantothenic acid is an integral precursor involved in the synthesis of coenzyme A (CoA) and functions in the decarboxylation of pyruvate and lipid metabolism in the citric acid cycle (62). Although multiple lines of evidence support these inferences, novel systematic approaches are needed to conclusively annotate unknown metabolite peaks.

Meta-analyzed and race-stratified GWAS in CARDIA mapped numerous overlapping SNP-metabolite peak associations in known loci of the human metabolome, suggesting a broad generalizability of associations across populations. We focused these analyses on only in-house library matches due to the higher evidence-basis for these matches. We found that our results largely overlapped with known associations and our novel SNP-metabolite pairs were well supported by known functions of nearby candidate genes. For example, variants in the *ALMS1/NAT8/ALMPS1* locus are primarily associated with *N-*acetylated amino acids, as *NAT8* shows amino acid sequence homology to *N*-acetyltransferases (63). Similarly, the *OPLAH* locus functions within the glutathione salvage pathway catalyzing the cleavage of 5-oxoproline to glutamate (64), thus differences in 5-oxoproline levels are expected. We did identify several novel locus-metabolite pairs with no clear link to gene function, including the *ALMPS1-*Phenylethanolamine and the *PM20D2*-Deoxyribose in the CARDIA meta-analyzed strata and the *SLC17A1*-Furan-2,5-dicarboxylic acid pair in the CARDIA White stratum. However, this does not necessarily mean that these associations are false as the breadth and diversity of the human metabolome is vast and the known functions of metabolites are constantly expanding. While metabolite reference libraries contain large numbers of compounds, matching signals to metabolites remains a challenging task, with many signals having low evidence matches (e.g., exact mass match only) or no matches at all. Indeed, these unmatched signals – termed the “dark matter” of metabolomics – may hold key information in mGWAS investigations and other metabolomics studies (65). Therefore, these associations which contain unannotated signals (or signals with low evidence matches) may need further profiling to establish their chemical substructure or compound similarities to other metabolite features to help clarify the role of metabolism in genomic association studies.

While we have shown there is a broad overlap of SNP-metabolite associations across race-stratified CARDIA analyses, recent metabolomic studies that focused on diverse populations have shown unique associations that were more common in Black (15) and Hispanic/Latino populations (1) suggesting population specific genetic architecture. Here, we found three population specific variants (rs466067 in *AGXT2*, rs558946049 in *SLC22A4* and rs79530723 near *SLC28A1*) among the Black CARDIA stratum associated with three different unannotated metabolite peaks. Two of these metabolite peaks were also associated with SNPs in the White CARDIA stratum, albeit with different sentinel variants (rs40199 in *AGXT2* and rs1939769 in *SLC22A4*). Only the rs79530723 variant near *SLC28A1* was specific to the Black stratum. Interestingly rs79530723 was previously associated with cytidine in the Hispanic Community Health Study/Study of Latinos (HCHS/SOL) cohort, likely due to the well-known African ancestry observed in Hispanic/Latino populations (1). Studies have shown *SLC28A1* encodes the concentrative nucleotide transporter 1 (CNT1) which is involved in cytidine and uridine transmembrane transporter activity (66). Furthermore, variants within *SLC28A1* and knock out mice models have also been associated with differences of serum metabolite levels especially N4-acetylcytidine measurements (6) and differences in the renal salvage of pyrimidine nucleosides; all of which support the *SLC28A1*-cytidine association in this study.

There are several limitations associated with this work. As is common in metabolomics studies (65), the majority of metabolomic peaks did not match the in-house library, limiting our ability to identify and interpret SNP-metabolite associations. In addition, mapping SNPs to genes in loci is still imprecise and as more metabolomic peaks are identified/annotated with improved technological advances, novel and more precise genetic signals will be found. As more metabolite peaks are matched to in-house libraries, mGWAS may reveal mechanistic insights for precision medicine. Despite these limitations, we identified an unreported metabolite peak – locus association which showed a functional connection with the citric acid cycle and identified one population-specific SNP-unannotated metabolite peak association in our CARDIA Black stratum. In addition, our study demonstrated consistency in the genetic architecture of circulating metabolites across populations. The insights gained from this study demonstrate the importance of studying diverse study populations to advance the science of the genetic underpinnings of the human metabolome.

## Data Availability

All data produced are available online at https://www.cardia.dopm.uab.edu/

## LIST of ABBREVIATIONS

NHW: Non-Hispanic White
CARDIA: Coronary Artery Risk Development in Young Adults
GWAS: Genome-wide Association Analysis Study
PheWAS: Phenome-wide Association Analysis Study
UHPLC: Ultra-high Performance Liquid Chromatography
ASCVD: Atherosclerotic Cardiovascular Disease
QCSPs: Quality Control Study Pools
LC-MS: Liquid Chromatography-Mass Spectrometry
NIST: National Institute of Standards and Technology
ADAP: Automated Data Analysis Pipeline
ANOVA: Analysis of Variance
ADAP-KDB: Analysis Pipeline Spectral Knowledgebase
RT: Retention Time
MS: Mass Spectrometry
MS/MS: Tandem Mass Spectrometry
HMDB: Human Metabolome Database
KEGG: Kyoto Encyclopedia of Genes and Genomes
CARe: Candidate Gene Association Resource
NHLBI: National Heart, Lung, and Blood Institute
MAF: Minor Allele Frequency
PC: Principal Components
SNP: Single-nucleotide Polymorphism
COJO: Conditional and Joint
GCTA: Genome-wide Complex Trait Analysis
VEP: Variant Effect Predictor
FET: Fisher’s Exact TTests
CMDKP: Common Metabolic Disease Knowledge Portal
KHNYN/SDR39U1: KH and NYN containing /Short Chain Dehydrogenase Reductase Family 39U Member 1
TCA: Citric Acid Cycle
NFE: non-Finnish European
HCHS/SOL: Hispanic Community Health Study/Study of Latinos

## DECLARATIONS

## Ethics approval and consent to participate

The study was conducted in accordance with the ethical standards of the Institutional Review Board at each CARDIA site and with the Declaration of Helsinki 1964 and its later amendments or comparable ethical standards and approved by the Institutional Review Board at the University of North Carolina at Chapel Hill and the CARDIA Steering Committee. This submission was reviewed by the Office of Human Research Ethics, which has determined that this submission does not constitute human subjects research as defined under federal regulations [45 CFR 46.102 (d or f) and 21 CFR 56.102(c)(e)(l)] and does not require IRB approval. Date: 7 December 2018.

## Consent for publication

Not applicable

## Availability of data and materials

The dataset supporting the conclusions of this article is available in the Coronary Artery Risk Development in Young Adults (CARDIA) study repository, https://www.cardia.dopm.uab.edu/.

## Funding and Acknowledgements

The authors are grateful for the support from the National Heart, Lung, and Blood Institute under R0HL143885. The Coronary Artery Risk Development in Young Adults Study (CARDIA) is conducted and supported by the National Heart, Lung, and Blood Institute (NHLBI) in collaboration with the University of Alabama at Birmingham (75N92023D00002 & 75N92023D00005), Northwestern University (75N92023D00004), University of Minnesota (75N92023D00006), and Kaiser Foundation Research Institute (75N92023D00003). This manuscript has been reviewed by CARDIA for scientific content.

## Authors’ contributions

MK, BRR, CLA and KEN conceived the study question. PGL, CLA, KEN and JPR obtained study funding. CEL collected samples. BRR, XD, SJS, SLM performed raw metabolomic profiling and data preprocessing. HMH and MG conducted genetic imputation. MK, MG, HMH, YT and KEN built the dataset and advised on genetic analyses. MK, BRR, KAM, AGH, VLB, SO, SL, CLA, PGL and KEN contributed to data research and interpretation. MK carried out the primary analysis and drafted the manuscript. All authors revised the manuscript critically for intellectual content, approved the version to be published, and agreed to be accountable for all aspects of the work.

## Disclaimer

The views expressed in this manuscript are those of the authors and do not necessarily represent the views of the National Heart, Lung, and Blood Institute; the National Institutes of Health; or the U.S. Department of Health and Human Services.

