## Supplementary Figures for "Integration of metabolomic and genetic data reveals novel variants underpinning the human metabolome: the Coronary Artery Risk Development in Young Adults (CARDIA) study"

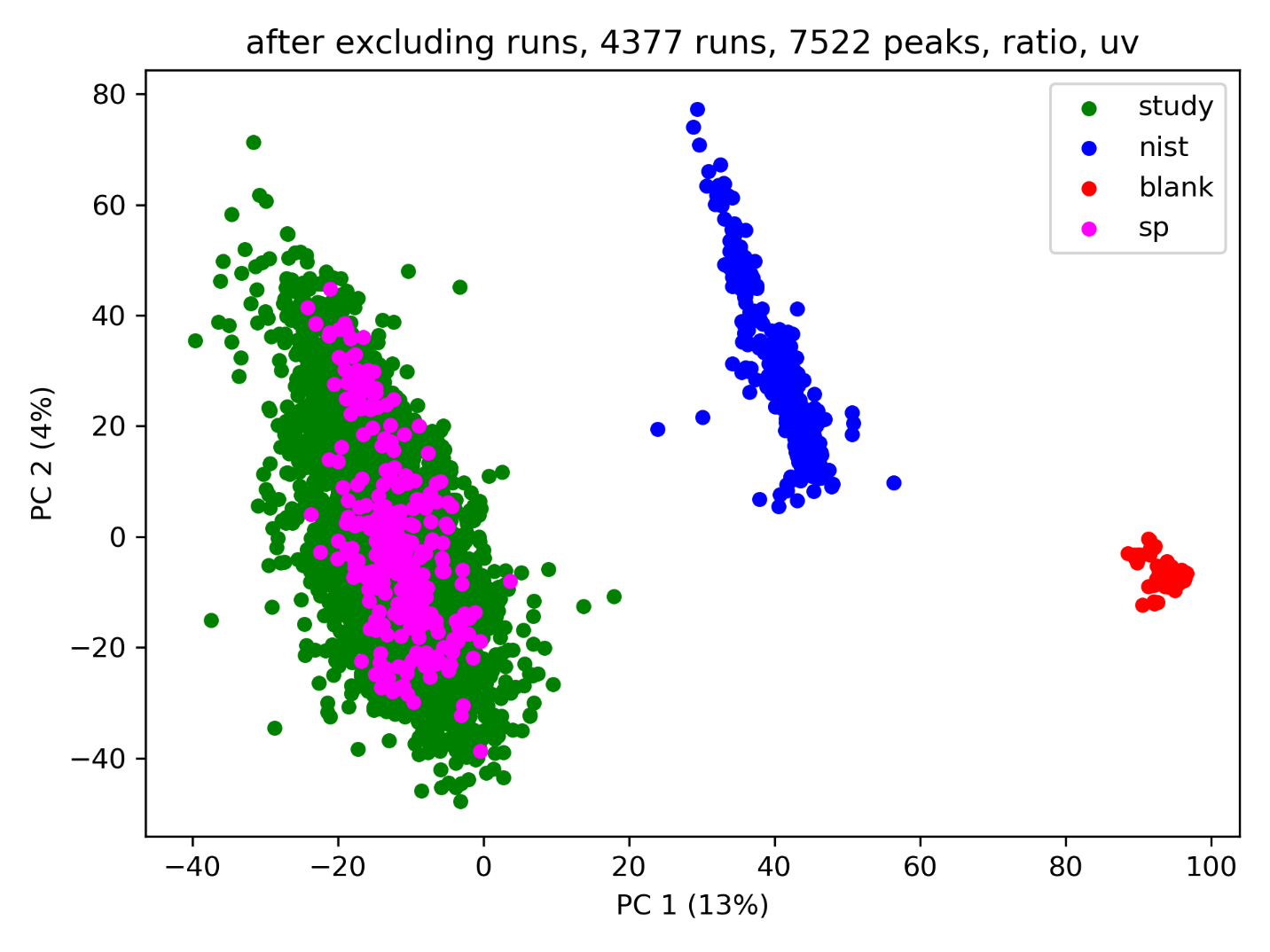
**Figure S1:** PCA of all study samples, study pools, NIST reference samples and blanks using all 7,522 metabolomic features. Data were scaled to unit variance.

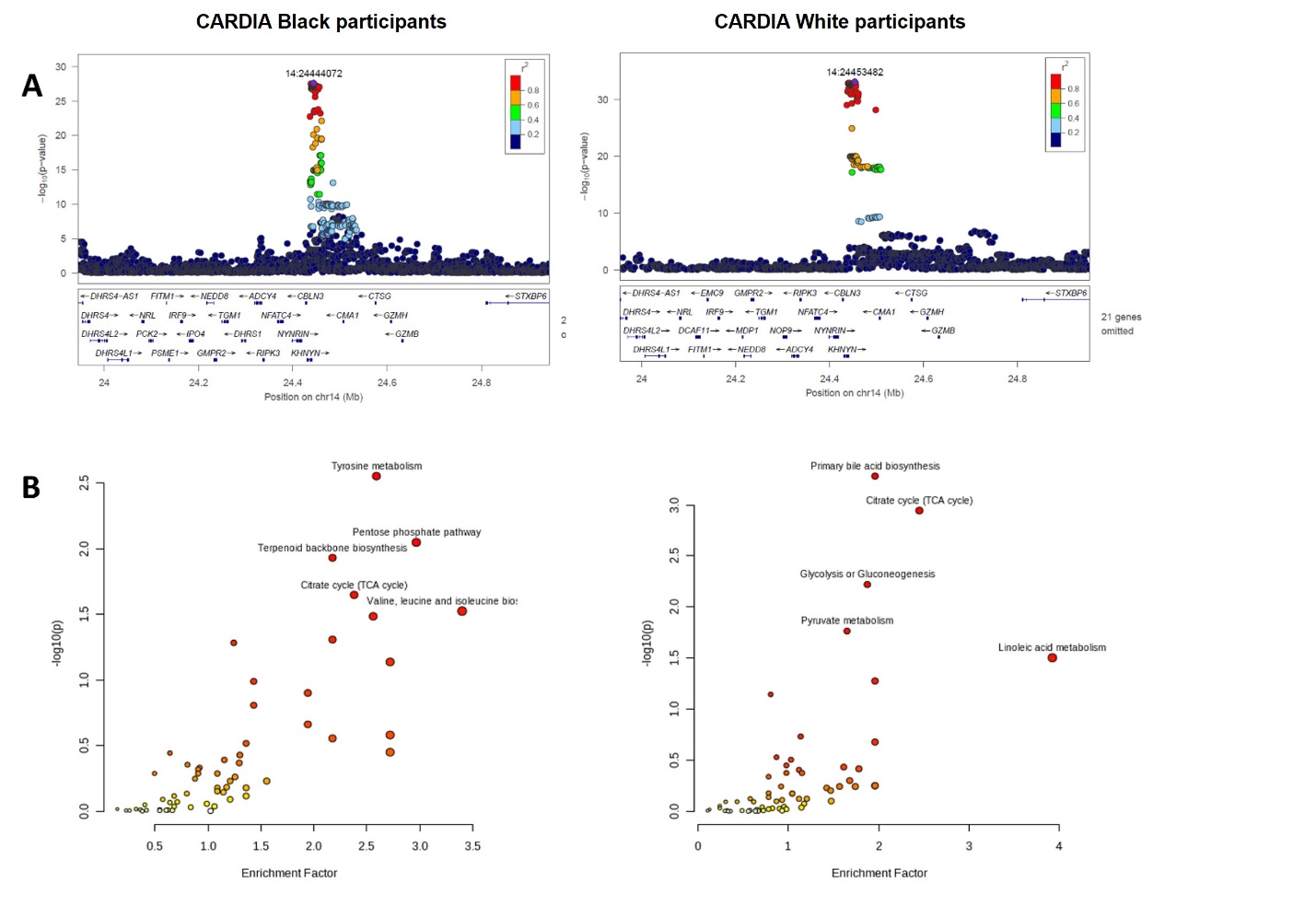

**Figure S2: A)** Regional association (LocusZoom) plot from the race-stratified analysis for the region encompassing *KHNYN/SDR39U1* in the CARDIA Black and White strata*.* The top variant is indicated with a purple diamond. The red line corresponds to a *P* value of 6.6 x10^-13^. The blue line corresponds to a *P* value of 5 x10^-8^. **B)** Dot plot of statistically significant pathways (<FET 0.05) from functional enrichment analysis of the CARDIA 7,522-metabolomic peaks using *P* values corresponding to the top variant in the CARDIA Black and White strata and *m/z* ratios for each peak. Each dot represents a metabolic pathway and is plotted according to its Fisher's exact test (FET) p-value on the x-axis. The dot size indicates the size of the enrichment factor, which represents the ratio of significant to total expected hits for each pathway.

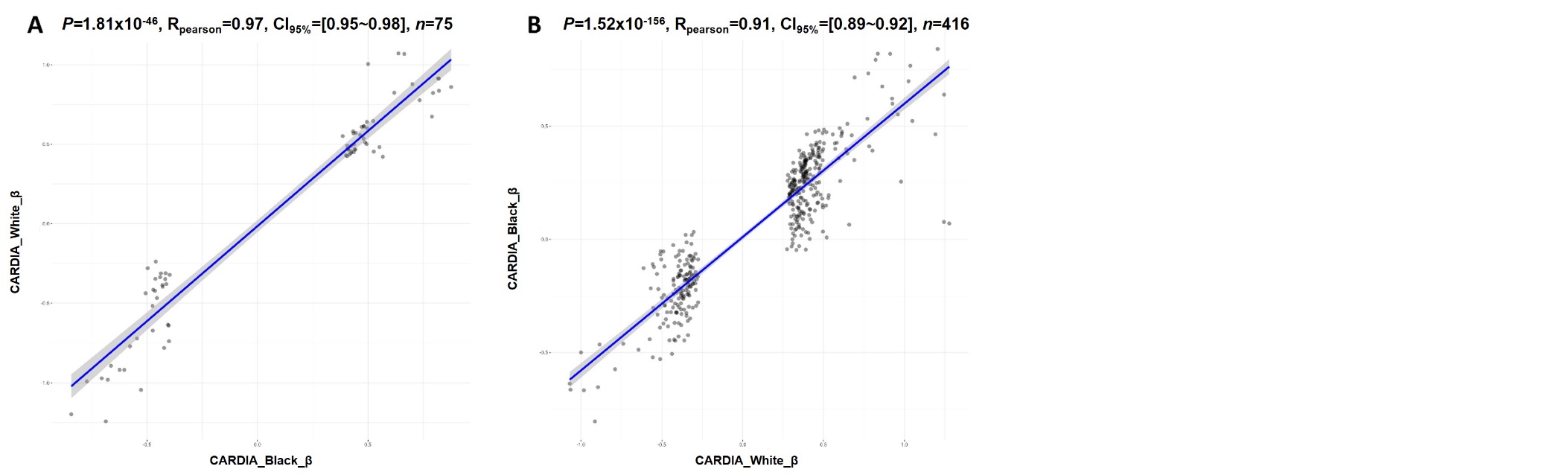

**Figure S3:** Distribution of variant based effects of metabolomic peaks in CARDIA **A)** between significantly mapped associations in the CARDIA Black stratum across the CARDIA White stratum and **B)** between significantly mapped associations in the CARDIA White stratum across the CARDIA Black stratum.
